# APOE Genotype and Fluid Biomarkers of Blood-Brain Barrier Integrity: A Systematic Review and Meta-Analysis

**DOI:** 10.64898/2026.09.27.26364123

**Authors:** Winnie Lucas, Melanie J. Murphy, Nina Riddell

## Abstract

Apolipoprotein E (APOE) genotype is theorised to influence dementia risk partly through effects on blood–brain barrier (BBB) function, but clinical studies examining APOE effects on BBB biomarkers report inconsistent results. Therefore, this systematic review and meta-analysis quantified associations between APOE genotype and two commonly used fluid biomarkers of CNS barrier function: cerebrospinal fluid (CSF)/serum albumin quotient (Q-Alb), and soluble platelet-derived growth factor receptor beta (sPDGFRβ). Ovid MEDLINE, Embase, and Web of Science were searched on 30^th^ June, 2026. Eligible peer-reviewed human studies reported Q-Alb or blood or CSF sPDGFRβ by APOE genotype. Risk of bias was assessed using the Joanna Briggs Institute checklist. Twenty-eight studies involving 5,375 participants were included: 17 assessed Q-Alb and 11 assessed sPDGFRβ. Random-effects meta-analyses determined that APOE ε2 carriage was associated with higher Q-Alb (*g* = 0.29, 95% CI 0.11 to 0.46). APOE ε4 carriage was not associated with Q-Alb overall (*g* = .13, 95% CI -.17 to .43) or sPDGFRβ overall (*g* = .10, 95% CI -.21 to .40). Genotype-specific Q-Alb analyses suggested medium positive but non-significant effects among ε4/ε4 and ε2/ε4 carrier subgroups. For sPDGFRβ, exploratory subgroup analysis revealed effect estimates near zero in cognitively unimpaired participants, and a small positive non-significant effect in cognitively impaired participants. Larger effects were also observed in samples with an older mean age, although this was not statistically significant. Overall, evidence for APOE ε4-associated alterations in Q-Alb and sPDGFRβ remained inconclusive, highlighting the need for further study across the spectrum of cognitive ageing and dementia. In contrast, APOE ε2 carriage was associated with greater barrier dysfunction as indexed by Q-Alb, broadening the predominant focus on APOE ε4 and identifying ε2 as a potentially important but understudied contributor to altered neurovascular function.

## 1. Introduction

Dementia affects an estimated 57.4 million people worldwide (Collaborators, 2022), of which Alzheimer’s disease (AD) represents the most common cause (Association, 2019). Neurovascular dysfunction is recognised as an early and central feature of dementia pathogenesis (Hattori, 2026; Sweeney et al., 2018), and apolipoprotein E (APOE) genotype may influence disease risk partly through effects on blood–brain barrier (BBB) function. The APOE ε4 allele, the strongest common genetic risk factor for late-onset AD and other dementias (Williams et al., 2026), has been proposed to promote BBB dysfunction through mechanisms including neuroinflammation and pericyte degeneration (Halliday et al., 2016; Nishitsuji, Kazuchika et al., 2011; Zlokovic, 2013). Fluid biomarkers related to BBB integrity have emerged as promising tools for monitoring neurovascular dysfunction (French et al., 2025) and may clarify genotype-specific neurovascular effects. However, studies comparing these biomarkers across APOE genotypes have produced inconsistent findings (e.g., Cicognola, C. et al., 2023; Janelidze et al., 2017; Libri et al., 2024; Riphagen et al., 2020), highlighting the need for further systematic quantitative synthesis.

Apolipoprotein E (ApoE) is a lipid-transport protein encoded by the APOE gene that delivers essential lipids and cholesterol to neurons, and is involved in clearing inflammatory components including amyloid-beta from the interstitial fluid (Troutwine et al., 2022). There are three major APOE alleles in humans: APOE ε2, APOE ε3, and APOE ε4, translating into ApoE2, ApoE3, and ApoE4 isoforms, respectively. The ε3 allele is the most common, accounting for approximately 80% of alleles, followed by ε4 and ε2 (Troutwine et al., 2022). Compositional differences between the APOE isoforms facilitate or impair receptor binding and lipid clearance capacity, resulting in different AD risk profiles (Phillips, 2014). Relative to the most common ε3 allele, AD risk is higher with ε4 carriage and lower with ε2 carriage (Williams et al., 2026). The effect of ε4 carriage is dose-dependent, with one ε4 allele conferring approximately 4.6-fold higher odds of AD diagnosis, and two ε4 alleles approximately 25-fold higher odds compared with non-ε4 carriers (Saddiki et al., 2020).

ApoE4 also increases risk for other dementias, including vascular, frontotemporal and Lewy body dementias, implicating this isoform in neurodegenerative pathology more broadly (Dickson et al., 2018; Koriath et al., 2019; Rohn, 2014; Tsuang et al., 2013). Indeed, the proportion of dementia burden attributable to APOE function is substantial, with recent analyses indicating that ε3 and ε4 carriage accounts for approximately half of all dementia cases and most AD cases (Williams et al., 2026).

Extensive preclinical evidence suggests that APOE ε4 promotes dementia pathogenesis through a multi-step pathway that includes effects on BBB integrity (Blumenfeld et al., 2024). The BBB is a dynamic neurovascular interface comprised of endothelial cells, sheathed by mural cells, including pericytes and vascular smooth muscle cells, and astrocytes. Tight junctions between endothelial cells form a highly selective barrier that regulates molecular transport between the circulation and the brain, maintaining central nervous system homeostasis (Sweeney et al., 2018). Astrocytes and vascular cells (endothelial cells, pericytes and smooth muscle cells) produce and secrete APOE (Blumenfeld et al., 2024). The ApoE4 isoform increases pro-inflammatory signalling in astrocytes (Arnaud et al., 2022; de Leeuw et al., 2022; Guo et al., 2004), increases pericyte mobility, proinflammatory signalling and degeneration (Bell et al., 2012; Casey et al., 2015; Halliday et al., 2016), and negatively impacts endothelial cell tight junctions and barrier formation (Nishitsuji, K. et al., 2011; Yamazaki et al., 2020), which can lead to BBB impairment (Bell et al., 2012; Halliday et al., 2016; Jackson et al., 2022) and allow blood-derived toxic molecules to enter the CNS (Bell et al., 2012). Interestingly, one recent human study suggested that the protective APOE ε2 allele may also modulate BBB permeability (Bernocchi et al., 2026).

APOE effects on BBB structure and function can be assessed using in vivo measures linked to BBB integrity, including neuroimaging and fluid biomarkers. While imaging techniques such as dynamic contrast-enhanced magnetic resonance imaging (DCE-MRI) provide spatial measures of BBB permeability, fluid biomarkers can provide insight into the biochemical processes underlying BBB dysfunction and offer a cost-effective and accessible approach to investigating BBB dysfunction that can be incorporated alongside established biomarker testing in research and clinical practice (French et al., 2025; Sun et al., 2021). The most widely used fluid biomarker of CNS barrier function is the CSF/serum albumin quotient (Q-Alb) (French et al., 2025; Sun et al., 2021). Because albumin is produced exclusively in the liver, its presence in the CNS reflects passive diffusion from the blood (Tumani and Hegen, 2015). Q-Alb corrects for individual differences in serum albumin concentrations, and this blood-CSF barrier (BCB) indicator is widely used as an indirect marker of global blood–brain/central nervous system barrier integrity (French et al., 2025; Reiber, 2003; Sun et al., 2021). Q-alb increases with age (Tibbling et al., 1977), is higher in males, and those with greater body mass index (Brettschneider et al., 2005; Parrado-Fernández et al., 2018; Skillback et al., 2021). It is also elevated in multiple types of dementia, particularly vascular dementia and dementia with Lewy bodies (Llorens et al., 2015; Musaeus et al., 2020; Skillbäck et al., 2017; Wong et al., 2022). In AD, meta-analyses have identified a small but significant Q-alb elevation (Olsson et al., 2016; Wang et al., 2026). It remains unclear whether Q-alb elevation is greater in the subset of AD patients who are APOE ε4 carriers, as results in studies examining the effect of genotype have been mixed (Janelidze et al., 2017; Libri et al., 2024; Padovani et al., 2025; Riphagen et al., 2020).

Q-alb is associated with protease and cytokine levels, apparently reflecting global inflammatory barrier disruption, rather than the focal vascular leakage detected by DCE-MRI (Hillmer et al., 2023), and thus elevation in dementia is consistent with evidence that immune-inflammatory responses and BBB breakdown contribute to neurodegeneration (Blumenfeld et al., 2024; French et al., 2025; Hattori, 2026; Sweeney et al., 2018). Given that ApoE4 is thought to promote BBB disruption partly through inflammatory mechanisms (Bell et al., 2012; Blumenfeld et al., 2024; Casey et al., 2015; Halliday et al., 2016), Q-Alb may theoretically be informative for detecting APOE ε4-associated BBB compromise. Therefore, further synthesis of the existing literature examining APOE genotype effects on Q-alb is warranted.

As discussed above, pericytes are critical for maintaining BBB integrity and are theorised to be a primary target of APOE ε4-mediated BBB dysfunction, with ε4 carriers showing greater pericyte degeneration on autopsy (Halliday et al., 2016). Such pericyte injury results in shedding of soluble platelet-derived growth factor receptor β (sPDGFRβ) into biofluids including CSF (Sagare et al., 2015). CSF sPDGFRβ levels correlate with DCE-MRI measures of BBB permeability in the hippocampus and parahippocampal gyrus (Montagne et al., 2020; Nation et al., 2019), and elevated sPDGFRβ in APOE ε4 carriers has been linked to inflammatory pericyte BBB breakdown pathways and predicts subsequent cognitive decline (Montagne et al., 2020). sPDGFRβ is correlated with Q-alb in most participant groups (Cicognola, Claudia et al., 2023; Lv et al., 2023; Miners et al., 2019; Nation et al., 2019), and like Q-alb, sPDGFRβ levels increase with age and are associated with inflammatory markers (Cicognola, Claudia et al., 2023; Preis, Lukas et al., 2024). As such, sPDGFRβ provides a complementary marker more closely linked to pericyte injury than to Q-Alb, though findings to date examining the effects of APOE genotype on sPDGFRβ levels have been mixed (Cicognola, Claudia et al., 2023; Lv et al., 2023; Montagne et al., 2020; Preis, Lukas et al., 2024; Sagare et al., 2026), potentially reflecting the varying age and disease states of participants.

As therapeutic strategies targeting the blood-brain barrier continue to emerge (Montagne et al., 2020; Searson and Banks, 2026), identifying individuals most likely to benefit from these interventions will require a better understanding of APOE-associated BBB dysfunction and the ability of fluid biomarkers to detect it. However, evidence linking APOE genotype to fluid biomarkers of BBB integrity remains inconsistent and has not been systematically synthesised. Therefore, this systematic review and meta-analysis aimed to quantify associations between APOE genotype and Q-Alb and sPDGFRβ biomarkers. Secondary analyses explored whether associations differed according to diagnostic status where feasible.

## 2. Methods

### 2.1 Search Strategy

This systematic review and meta-analysis was conducted according to the Preferred Reporting Items for Systematic Reviews and Meta-Analyses (PRISMA) framework (Page et al., 2021), and was pre-registered via the Open Science Framework (https://doi.org/10.17605/OSF.IO/YX52P). Final literature searches were conducted on 30^th^ June 2026 across Ovid MEDLINE, Web of Science, and Embase, with additional manual searching via Google Scholar. This combination of databases was selected for its excellent recall rate of health-related literature (Bramer et al., 2017). Key terms included words related to Apolipoprotein E, albumin quotient, and sPDGFRβ, with Boolean operators and MeSH headings to capture variations and associated terms. Table 1 depicts the search strategy formulated for Ovid MEDLINE.

**Table 1.** Search Strategy in Ovid MEDLINE.

| Row | Search command |
| --- | --- |
| 1. | Apolipoprotein E/ |
| 2. | APOE*.mp |
| 3. | 1 or 2 |
| 4. | Albumin.mp |
| 5. | Albumin quotient.mp |
| 6. | Qalb.mp |
| 7. | Q-alb.mp |
| 8. | Albumin ratio.mp |
| 9. | CSF/serum albumin.mp |
| 10. | Platelet derived growth factor* |
| 11 | PDGF*.mp |
| 12. | Soluble platelet derived growth factor* |
| 13. | sPDGFR*.mp |
| 14. | 4 or 5 or 6 or 7 or 8 or 9 or 10 or 11 or 12 or 13 |
| 15. | 3 and 14 |
| 16 | Limit 15 to English and humans only |

### 2.2 Study Selection Criteria

Search results were imported into Covidence (Innovation, 2026), where duplicates were automatically and manually identified and removed. Studies were screened in Covidence by two independent reviewers. Conflicts were resolved through discussion with a third independent reviewer to achieve consensus. During the first stage of screening, reviewers read the titles and abstracts of each study to assess whether they met the selection criteria. In the second stage of screening, all relevant texts were read in their entirety by two independent reviewers and assessed against a hierarchy of exclusion criteria (Table 2). Peer-reviewed, English language, original studies reporting CSF/serum Q-Alb or blood or CSF sPDGFRβ BBB integrity measures in APOE genotyped human participants were eligible for inclusion. Studies could include participants with AD or other dementias, other health conditions, or healthy controls. Reviews, case studies and case series were excluded, as were animal and in vitro studies.

**Table 2.** Exclusion Criteria Hierarchy.

| Level | Exclusion criteria |
| --- | --- |
| 1. | Full text not available |
| 2. | Study not in English |
| 3. | Study not examining human participants |
| 4. | APOE genotype categories not reported |
| 5. | Plasma/CSF sPDGFRβ or Q-Alb not reported |
| 6. | Study not reporting Q-Alb/sPDGFRβ results stratified by APOE genotype |

To enable extraction of relevant data, participants were required to be stratified by APOE ε4 status or specific APOE genotype, and APOE ε4 status groups or APOE genotypes compared with respect to Q-Alb values or sPDGFRβ concentrations. Where studies included the relevant variables but did not stratify BBB marker results by APOE genotype or did not report individual datapoints or means and standard deviations (SD) (or standard errors (SE)) for the stratified comparisons, study authors were contacted to request the relevant data. If data were not provided the study was either excluded where genotype stratified results were not available, or included in the narrative synthesis where stratified results were available but were unable to be meta-analysed.

### 2.3 Quality Assessment

The Joanna Briggs Institute Critical Appraisal Tool’s Checklist for Analytical Cross Sectional Studies (Moola et al., 2020) was used to assess risk of bias. The original checklist was adapted to accommodate the study topic, with eight items included in the checklist against which each study was evaluated: 1) clarity of inclusion criteria, 2) clear definition of participants and setting, 3) reliability of APOE genotyping procedure, 4) use of objective, standard criteria for APOE genotype categorisation, 5) identification of confounding factors, 6) identification of strategies to deal with confounding factors, 7) reliability of outcome (Q-Alb/sPDGFRβ) measurement, and 8) use of appropriate statistical analyses. Ratings were ‘low’ (low risk of bias), ‘unclear’ (unable to determine risk of bias), or ‘high’ (high risk of bias). Risk of bias assessment outcomes were visualised using robvis (McGuinness and Higgins, 2021).

### 2.4. Data Extraction and Analysis

Data from studies that progressed through full text screening were manually extracted. As shown in Table 3, publication details, APOE genotype, and Q-Alb and/or sPDGFRβ measurement details were extracted. Participant characteristics that could impact BBB integrity measures including age, sex, and diagnostic status, were also extracted. When allelic stratification was reported, the effect sizes of each allelic group were extracted separately to facilitate both overall and subgroup analyses.

**Table 3.**
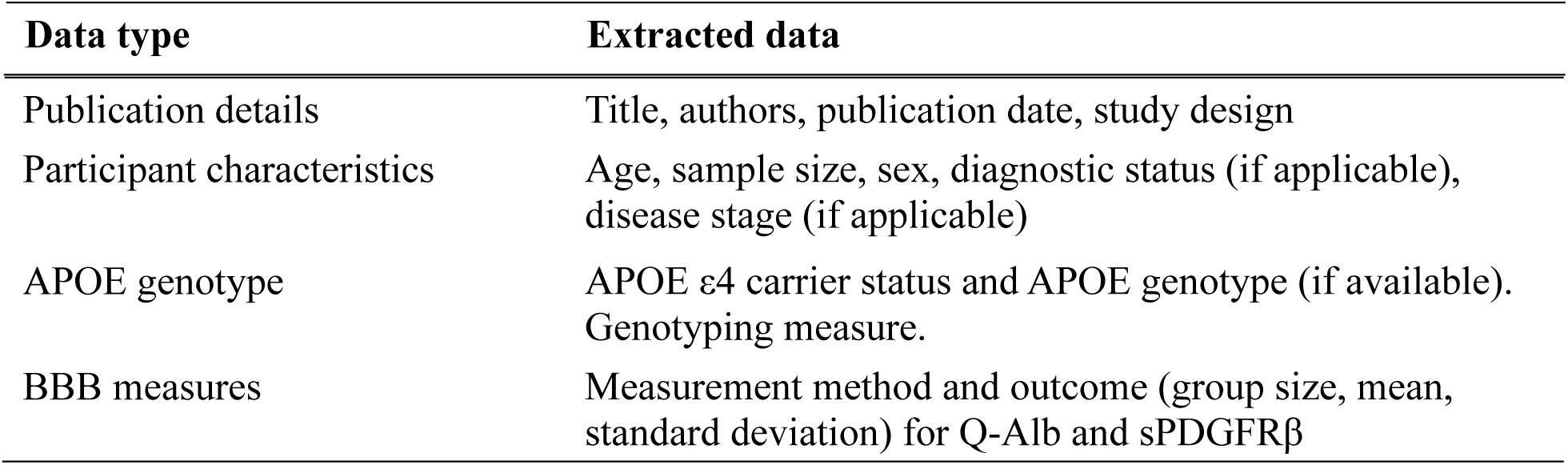
Data extracted from included studies.

| **Data type** | **Extracted data** |
| --- | --- |
| Publication details | Title, authors, publication date, study design |
| Participant characteristics | Age, sample size, sex, diagnostic status (if applicable), disease stage (if applicable) |
| APOE genotype | APOE ε4 carrier status and APOE genotype (if available). Genotyping measure. |
| BBB measures | Measurement method and outcome (group size, mean, standard deviation) for Q-Alb and sPDGFRβ |

Where necessary, means and SDs were pooled to create one mean and SD for each genotype grouping (e.g., combining E2/E3 and E3/E3 into an APOE ε4 non-carriers group) (Arsham, 2015; Chandler et al., 2019). When data were present only in a figure, PlotDigitizer Version 3 (Aydin and Yassikaya, 2022) was used to convert visual datapoints into numbers for extraction into Excel. When 95% confidence intervals were reported instead of SDs, these values were converted into SDs using Meta-Analysis Accelerator (Abbas et al., 2024).

Statistical analyses were conducted using JASP Version 0.98.1 (2026). The primary analyses compared Q-Alb and sPDGFRβ levels between APOE ε4 carriers and non-carriers. Following evidence of a larger Q-Alb effect among ε2/ε4 carriers and emerging literature implicating APOE ε2 in BBB function (Bernocchi et al., 2026), an exploratory analysis examined Q-Alb differences between APOE ε2 carriers and non-carriers. For Q-Alb, several studies contributed multiple non-independent effect sizes because the same control/comparator group was used for multiple allele comparisons, and studies also contributed effects across multiple diagnostic categories. To account for this dependency, four-level random-effects meta-analyses were conducted with effect sizes nested within samples and samples nested within studies. Between-study variance was estimated using restricted maximum likelihood (REML), and subgroup analyses were performed to examine differences according to APOE allelic subgroup and participant diagnosis.

For sPDGFRβ, the available data did not permit allelic subgroup analyses and each comparator group (i.e., APOE ε4 non-carrier group) contributed to a single effect size. Therefore, a single-level random-effects meta-analysis was performed using REML to examine the association between APOE ε4 carrier status and sPDGFRβ levels. Exploratory subgroup analyses examined cognitive/diagnostic status, and meta-regression examined mean participant age as a potential moderator. The Knapp–Hartung adjustment was applied to improve type I error control for random-effects meta-analyses with a small to moderate number of studies and substantial between-study heterogeneity (Jackson et al., 2017).

For both Q-Alb and sPDGFRβ analyses, Hedges’ *g* was used as the effect size metric to quantify standardised mean differences between APOE ε4 or ε2 carriers and non-carriers (Borenstein et al., 2021). Pooled effect estimates and 95% confidence intervals were calculated for each analysis and displayed using forest plots. Effect sizes of 0.2, 0.5, and 0.8 were interpreted as small, medium, and large, respectively (Cohen, 2013; Gallardo-Gomez et al., 2024). Statistical heterogeneity was assessed using Cochran’s *Q* statistic, with a significant result (*p* < .05) indicating heterogeneity among studies (Cochran, 1954). Between-study heterogeneity was quantified using τ². For analyses including at least 10 studies, publication bias and small-study effects were assessed by visual inspection of funnel plots (Supplementary Figures 1-2) and Egger’s weighted regression test (Hu et al., 2026). Eligible studies that could not be quantitatively synthesised were summarised narratively.

When data permitted, sensitivity analysis was conducted to examine the robustness of pooled estimates. These included leave-one-study-out analyses, exclusion of studies where risk of bias assessment suggested potential confounding by age or sex, exclusion of studies with samples containing a mix of disease categories, or a mix of alleles (i.e., APOE4+ versus APOE4-), and exclusion of studies that measured sPDGFRβ in blood samples (instead of CSF).

## 3. Results

### 3.1 Study Selection

Figure 1 presents a Preferred Reporting Items for Systematic Reviews and Meta-Analyses (PRISMA) flow diagram depicting the study selection process. Database searching identified 1600 abstracts (938 following removal of duplicates). Following abstract screening, 53 full texts were assessed, 24 of which were excluded for the following reasons: full text not available (1), APOE genotyping not performed (2), plasma/CSF sPDGFRβ or Q-Alb not measured (4), or necessary statistics not reported (17). Eighteen Q-Alb studies and 11 sPDGFRβ studies were eligible for inclusion.

**Figure 1.**
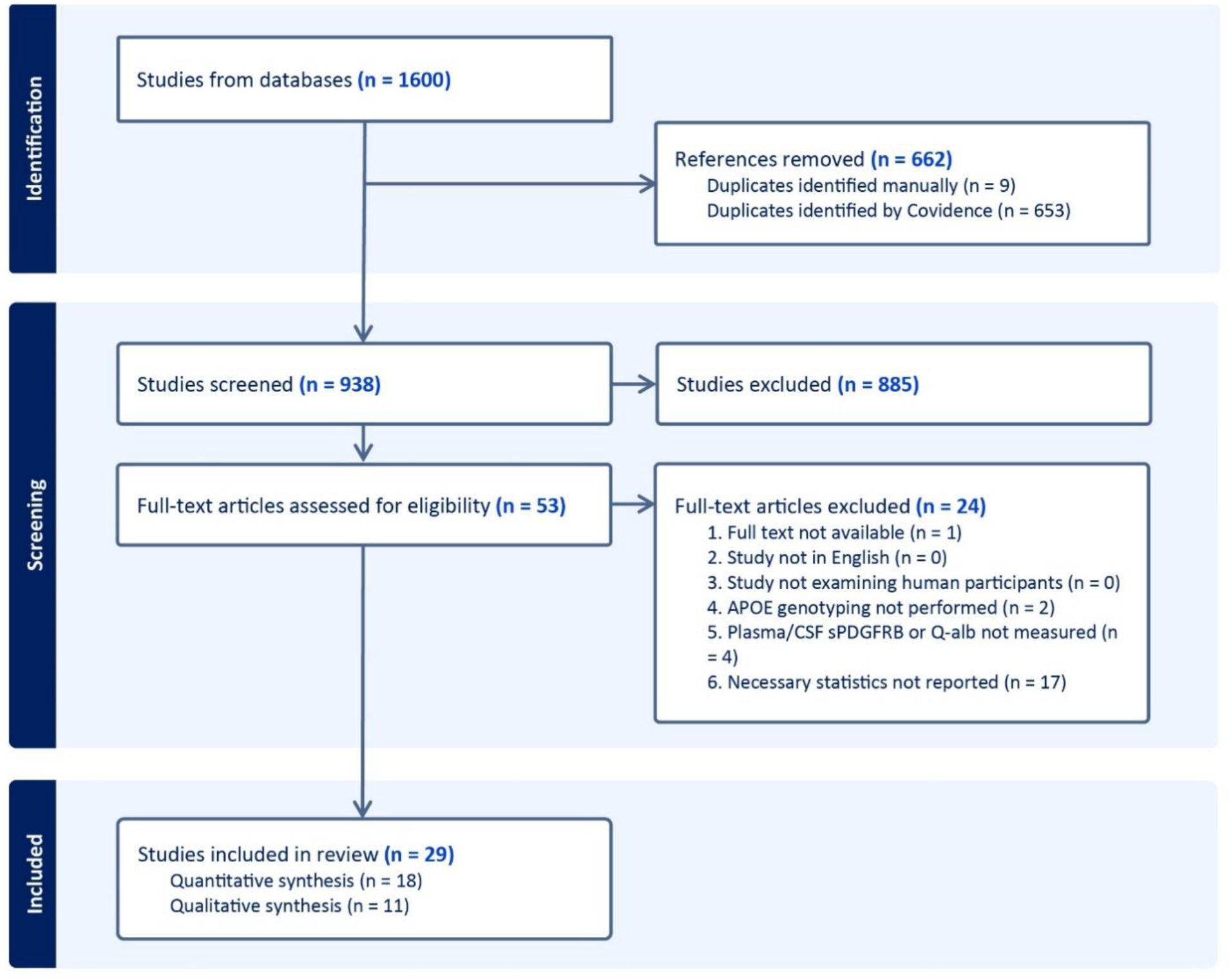
PRISMA flow diagram depicting the study selection process. One Q-Alb study was subsequently excluded during the data extraction phase, as outlined in the Quality Assessment and Risk of Bias section.

**Figure 2.**
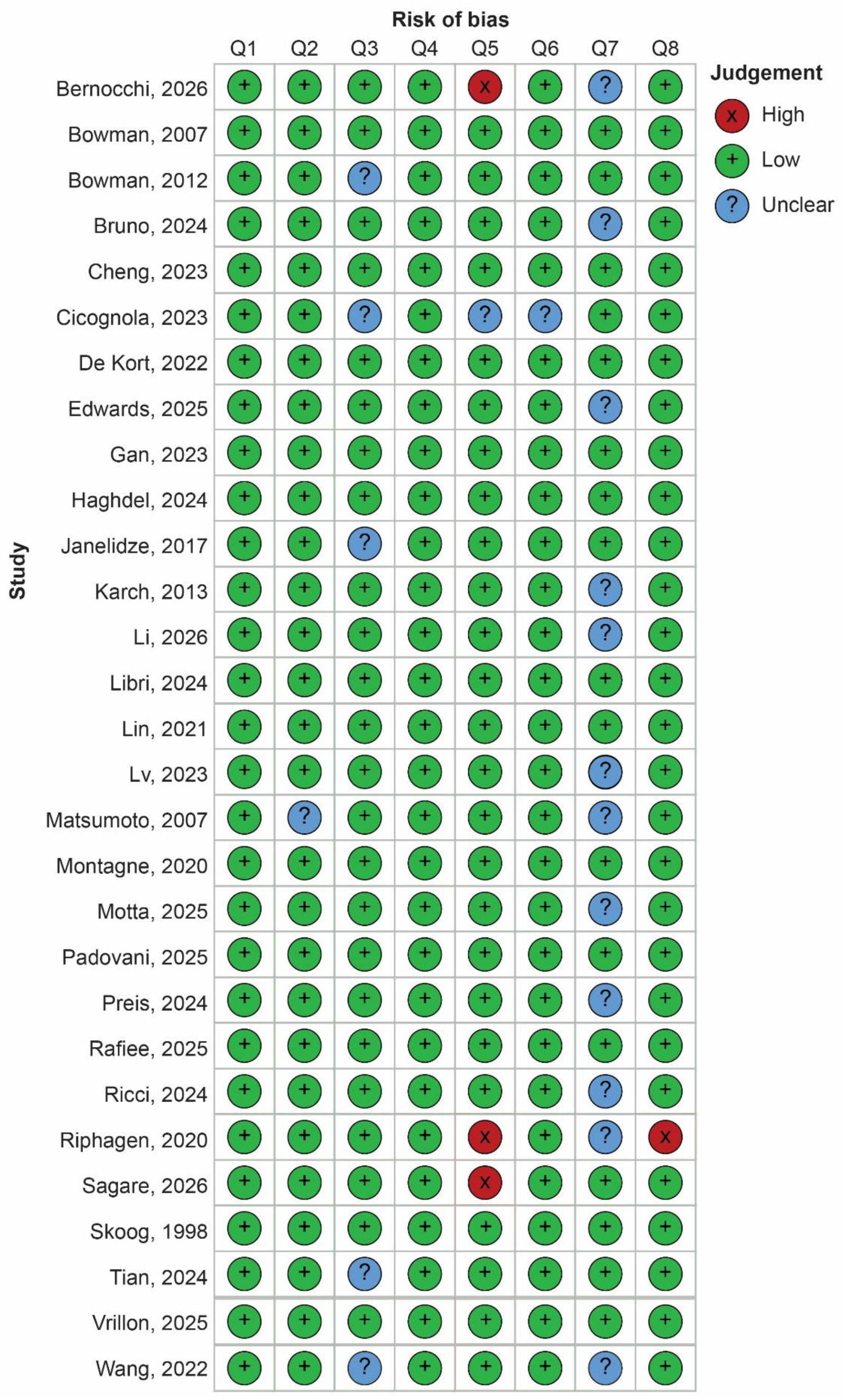
Risk-of-bias assessment of studies meeting inclusion criteria. Risk of bias was assessed using an adapted Joanna Briggs Institute Critical Appraisal Checklist for Analytical Cross-Sectional Studies. Traffic-light ratings indicate low (green), unclear / no information (blue), or high (red) risk of bias for each criterion. Q1, inclusion criteria; Q2, participants and setting; Q3, APOE genotype determination; Q4, genotype categorisation; Q5, identification of confounding factors; Q6, strategies to address confounding; Q7, outcome measurement; Q8, statistical analysis.

### 3.2 Quality Assessment and Risk of Bias

All studies clearly reported their inclusion criteria, and participant characteristics and study settings were adequately described, except for one study in which the recruitment setting was unclear (Matsumoto et al., 2007). Risk of bias for APOE genotype determination (Item 3) was rated unclear when the specific genotyping or phenotyping procedure was not reported (Bowman et al., 2012; Cicognola, Claudia et al., 2023; Janelidze et al., 2017; Tian et al., 2024). Outcome measurement (Item 7) was rated unclear when studies did not report the method used to measure albumin (Bernocchi et al., 2026; Bruno et al., 2024; Karch et al., 2013; Matsumoto et al., 2007; Motta et al., 2025; Ricci et al., 2024), did not adequately describe CSF and blood collection procedures (Riphagen et al., 2020), or did not report intra- and inter-assay coefficients of variation for sPDGFRβ ELISA measurements (Edwards et al., 2025; Li et al., 2026; Lv et al., 2023; Preis, L. et al., 2024), precluding assessment of assay precision. Four studies showed potential imbalances in age or sex between APOE groups (Bernocchi et al., 2026; Cicognola, Claudia et al., 2023; Riphagen et al., 2020; Sagare et al., 2026); three accounted for these potential confounders in subsequent analyses and were therefore rated at low risk of bias for confounding adjustment (Item 6). One study contained apparent data-entry errors in the table reporting data required for effect-size calculation and was therefore rated at high risk of bias for statistical analysis (Item 8). As the apparent errors prevented reliable extraction and interpretation of the relevant data, the study was excluded (Riphagen et al., 2020). Sensitivity analyses excluding the remaining studies with potential age or sex imbalances from meta-analyses did not materially alter the relevant pooled estimates (Supplementary Figures 3-5).

### 3.3 Q-Alb Study Characteristics

Supplementary Table 1 outlines the characteristics of studies examining the effects of APOE genotype on Q-Alb. Studies meeting inclusion and risk of bias criteria measured Q-Alb in a total of 3360 participants. The mean age of participants in these studies was between 62 and 85 and, with some exceptions (Bowman et al., 2007; Bowman et al., 2012; Ricci et al., 2024; Skoog et al., 1998), 50-65% of participants were female. Participants were primarily patients recruited from clinical settings such as hospitals or dementia care clinics, with almost all participants having a dementia diagnosis (primarily Alzheimer’s disease, with vascular dementia, dementia with Lewy bodies, frontotemporal dementia, and Creutzfeldt-Jakob disease also represented). Some studies included participants with mild cognitive impairment or subjective cognitive decline (Bruno et al., 2024; Janelidze et al., 2017; Libri et al., 2024; Lin et al., 2021; Rafiee et al., 2025; Tian et al., 2024), and three studies included cognitively unimpaired controls (Cheng et al., 2023; Lin et al., 2021; Rafiee et al., 2025).

### 3.4 Q-Alb Meta-Analysis

Analysis of all studies with quantitative data available for extraction revealed a negligible overall effect of APOE ε4 carriage on Q-alb (*g* = .13, 95% CI [-.17, .43]; Figure 3). Visual inspection of the funnel plot did not indicate marked asymmetry (Supplementary Figure 1), and Egger’s weighted regression test did not provide evidence of small-study effects (*t* = 1.492, *p* = .147). When studies with mixed allele categories (i.e., comparing ε4 carriers to non-carriers) were removed in sensitivity analyses, the overall effect size was small to medium and non-significant (*g* = .38, 95% CI [-0.52, 1.28]; Supplementary Figure 5).

**Figure 3.**
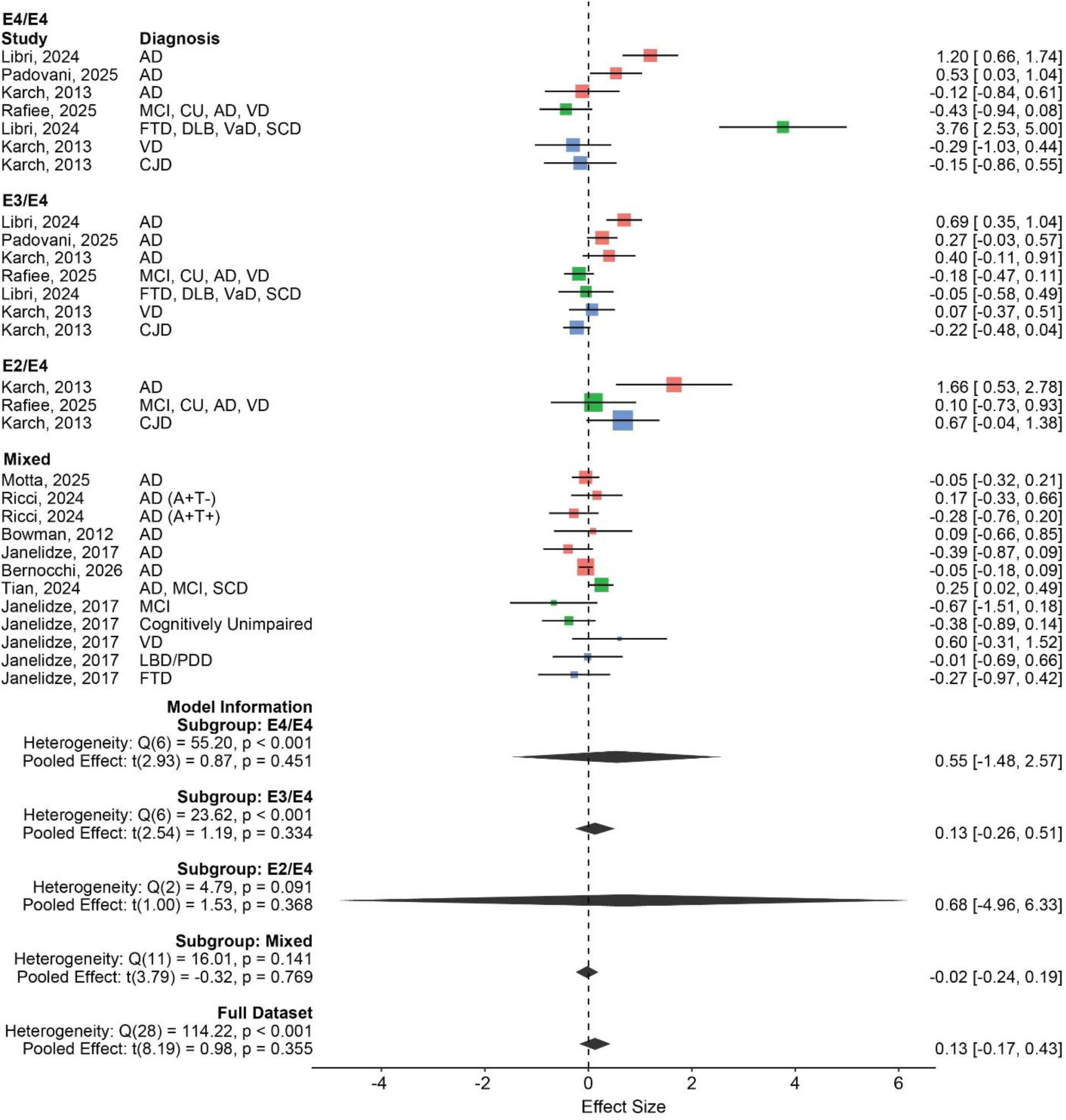
Forest plot of the association between APOE ε4 carrier status and Q-Alb levels. Squares represent study-specific Hedges’ g effect sizes, with square size proportional to study weight. Horizontal lines indicate 95% confidence intervals, and the diamond represents the pooled random-effects estimate. Effects are coloured according to diagnosis category (green = mixed sample, blue = non-AD dementia, red = AD). Positive effect sizes indicate higher Q-Alb levels in APOE ε4 carriers, whereas negative effect sizes indicate higher levels in non-carriers. In Ricci et al., (2024), A+T+ refers to patients with amyloid-beta and tau pathology. A+T-refers to patients with amyloid-beta pathology, but not tau pathology.

To investigate whether the association between APOE ε4 carrier status and Q-Alb differed according to allelic variation, subgroup analyses compared Q-Alb levels in APOE ε2/ ε 4, APOE ε3/ε4, APOE ε4/ε4, and mixed APOE ε4 carrier groups (where the specific allelic combination was not reported) relative to non-carriers. Medium positive pooled effects were observed for Q-Alb among homozygous APOE ε4/ε4 carriers (*g* = 0.55, 95% CI [−1.48, 2.57]) and APOE ε2/ε4 carriers (*g* = 0.68, 95% CI [−4.96, 6.33]). However, neither effect was statistically significant, and the extremely wide confidence intervals indicate substantial uncertainty. Sensitivity analyses further indicated that the study by Libri et al. (2024) strongly influenced the pooled effect for APOE ε4/ε4 carriers (Supplementary Figure 3). For APOE ε3/ε4 carriers (*g* = 0.13, 95% CI [−0.26, 0.51]) and mixed APOE ε4 carrier groups (*g* = −0.02, 95% CI [−0.24, 0.19]), in which the specific allelic combination was not reported, effect sizes were negligible to small (Figure 3). Further subgroup analysis comparing AD, non-AD dementia, and mixed sample groups, provided no evidence that the association between APOE ε4 carrier status and Q-Alb differed across these diagnostic categories (Supplementary Figure 10).

Given the medium-to-large effect observed among APOE ε2/ε4 carriers, together with emerging evidence implicating APOE ε2 in blood–brain barrier permeability, additional exploratory analyses were conducted to examine the association between APOE ε2 carriage and Q-Alb. Across all included studies, APOE ε2 carriage was associated with a small but statistically significant increase in Q-Alb (*g* = 0.29, 95% CI [0.11, 0.46]; Figure 4).

**Figure 4.**
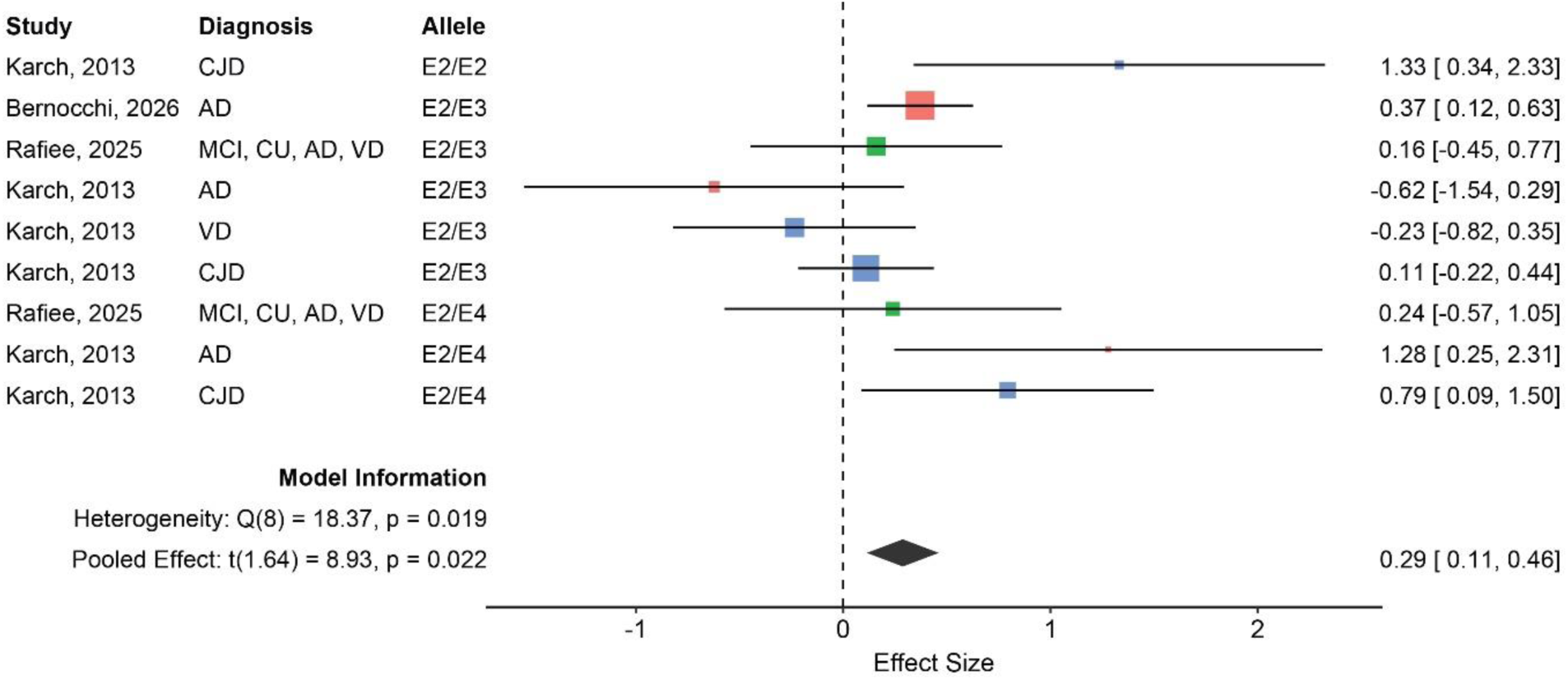
Forest plot of the association between APOE ε2 carrier status and Q-Alb levels. Squares represent study-specific Hedges’ g effect sizes, with square size proportional to study weight. Horizontal lines indicate 95% confidence intervals, and the diamond represents the pooled random effects estimate. Effects are coloured according to diagnosis category (green = mixed sample, blue = non-AD dementia, red = AD). Positive effect sizes indicate higher Q-Alb levels in APOE ε2 carriers, whereas negative effect sizes indicate higher levels in non-carriers. Additional subgroup analysis exploring the impact of allelic variations on this effect are presented in the supplementary materials. As only one effect was available for APOE ε2/ε2 carriers, this subgroup analysis compares APOE ε2/ε3 and APOE ε2/ε4 carrier groups only.

Heterogeneity was significant (*Q*(8) = 18.37, *p* = 0.019). Further exploratory subgroup analyses suggested a larger effect among APOE ε2/ε4 carriers (*g* = .65, 95% CI [-3.89, 5.20]) than among APOE ε2/ε3 carriers (*g* = .17, 95% CI [-.66, 1.01]; Supplementary Figure 11).

However, these estimates were based on few studies and were imprecise, particularly for the APOE ε2/ε4 subgroup, and should therefore be interpreted cautiously.

### 3.5 Q-Alb Narrative Synthesis

Key findings from an additional seven studies examining the association between APOE ε4 carriage and Q-Alb that could not be included in the quantitative synthesis are summarised in Supplementary Table 2. Consistent with the pooled analysis, these studies reported no statistically significant association between APOE ε4 carriage and Q-Alb in mixed APOE ε4 carrier groups. Skoog et al. additionally examined allele-specific carriage and reported higher Q-Alb among APOE ε3 non-carriers (i.e., ε2/ε2, ε2/ε4, or ε4/ε4 genotypes), consistent with the meta-analytic findings, but found no significant differences for ε4 or ε2 carriage versus non-carriage.

### 3.6 sPDGFRβ Study Characteristics

Supplementary Table 3 outlines the characteristics of studies examining the effects of APOE genotype on sPDGFRβ. Studies meeting inclusion and risk of bias criteria measured sPDGFRβ in a total of 2,015 participants. The mean age of participants was similar to that of Q-Alb studies (60 – 74), and female participant representation ranged from 42% – 86%. sPDGFRβ participants were typically recruited through university research centres, were cognitively unimpaired (> 50%; 9/11 studies), reported subjective cognitive decline (1/11) or had a diagnosis of mild cognitive impairment (10/11 studies) or dementia (primarily AD; 6/11 studies). One study assessed participants with Cerebral Amyloid Angiopathy (CAA). This diagnostic profile differs from that of participants in Q-Alb studies where most participants had a dementia diagnosis, and very few participants were cognitively unimpaired.

### 3.7 sPDGFRβ Meta-Analysis

Meta-analysis of sPDGFRβ data showed a negligible, non-significant positive association between APOE ε4 carriage and sPDGFRβ (*g* = .10, 95% CI [-.21, .40]; Figure 5). Exploratory subgroup analyses examined effects according to cognitive/diagnostic status. Pooled effects were close to zero among cognitively unimpaired participants (*g* = .04, 95% CI [-.88, .96]) and mixed diagnostic samples (*g* = -.01, 95% CI [-.34, .32]), whereas a small positive but non-significant effect was observed among cognitively impaired participants (*g* = .27, 95% CI [-.74, 1.29]). Substantial heterogeneity was present within the cognitively impaired subgroup (*Q*(3) = 14.19, *p* = .003; *I*² = 80.76%). Exploratory meta-regression further examined mean age as a potential moderator (Supplementary Figure 12). Across the full dataset, the predicted APOE ε4 effect increased with age (−1 SD: *g* = −0.17, 95% CI [−0.64, 0.30]; mean: *g* = 0.07, 95% CI [−0.22, 0.35]; +1 SD: *g* = 0.30, 95% CI [−0.10, 0.70]), but age was not a significant moderator (F(1, 8) = 2.66, *p* = 0.142). Substantial residual heterogeneity remained (*Q*(8) = 18.75, *p* = .016; *I*² = 65.75%).

**Figure 5.**
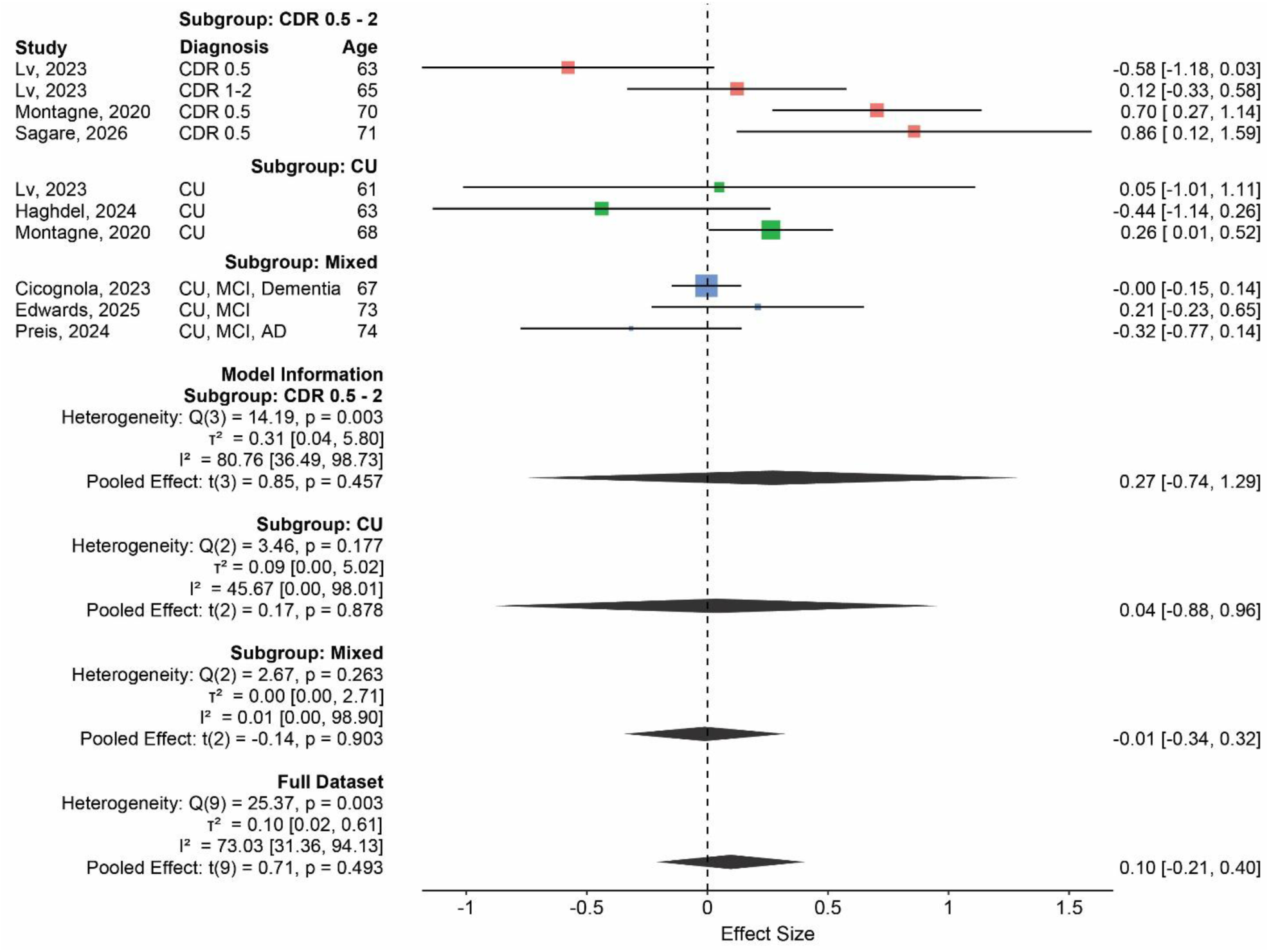
Meta-analysis of the association between APOE ε4 carrier status and sPDGFRβ levels. Squares represent study-specific Hedges’ g effect sizes, with square size proportional to study weight. Horizontal lines indicate 95% confidence intervals, and the diamond represents the pooled random-effects estimate. Effects are coloured according to diagnosis category (green = cognitively unimpaired, blue = mixed sample, red = Clinical Dementia Rating (CDR) score 0.5 – 2). Positive effect sizes indicate higher sPDGFRβ levels in APOE ε4 carriers, whereas negative effect sizes indicate higher levels in non-carriers.

### 3.8 sPDGFRβ Narrative Synthesis

Additional findings that could not be quantitatively synthesised provided complementary evidence regarding APOE genotype and sPDGFRβ. Vrillon et al (Vrillon et al., 2025), De Kort et al (De Kort et al., 2022), Wang et al (Wang et al., 2022) and Li et al (Li et al., 2026) found no evidence of an association between APOE ε4 carriage and CSF or plasma sPDGFRβ in CAA, MCI, pre-clinical AD, AD, dementia or mixed normal control, SCD and MCI groups. However, Vrillon et al found that APOE ε4 carriers had significantly lower CSF PDGFRβ levels in a small control sample (*N* = 23) (Vrillon et al., 2025). These findings are consistent with the negligible to small overall association between APOE ε4 carriage and sPDGFRβ observed in the quantitative synthesis.

Montagne et al. (Montagne et al., 2020), whose ε4 carrier versus non-carrier comparison was included in the meta-analysis, additionally reported no significant difference in CSF sPDGFRβ between carriers of one versus two ε4 alleles. Together with the null findings of De Kort et al (De Kort et al., 2022), who compared CAA APOE ε4 carriers versus non-carriers and ε2 carriers versus non-carriers, this provides little evidence for genotype-dose differences in CSF sPDGFRβ.

Finally, additional data from the cognitively unimpaired (CDR = 0) group reported by Sagare et al. (Sagare et al., 2026) could not be reliably extracted from the published figures. However, when considered alongside the cognitively impaired (CDR = 0.5) data included in the meta-analysis, the study reported higher sPDGFRβ among cognitively impaired APOE ε4 carriers than both cognitively unimpaired ε4 carriers and cognitively impaired ε3 homozygotes. In contrast, sPDGFRβ did not differ between cognitively unimpaired ε3 and ε4 groups. These findings are consistent with the findings of the subgroup analysis, in which APOE ε4 had a small effect on sPDGFRβ levels in cognitively impaired individuals, but a negligible effect in cognitively unimpaired and mixed participant groups. Collectively, the quantitative and narrative evidence therefore suggests that the association between APOE ε4 and sPDGFRβ may vary with cognitive or disease status, but the limited evidence currently available precludes firm conclusions.

## 4. Discussion

This systematic review and meta-analysis synthesised 28 studies comprising 5,375 participants examining associations between APOE genotype and fluid biomarkers linked to blood–brain barrier (BBB) integrity (French et al., 2025). The most robust finding was significantly higher Q-Alb among APOE ε2 carriers, suggesting greater blood-CNS barrier permeability. In contrast, APOE ε4 carriage was not significantly associated with Q-Alb overall. Genotype-specific analyses suggested medium elevations in Q-Alb among ε4/ε4 and ε2/ε4 carriers relative to ε4 non-carriers. However, these estimates were non-significant and highly imprecise. APOE ε4 carriage was associated with a negligible, non-significant elevation in sPDGFRβ. Exploratory subgroup analyses yielded a small non-significant positive effect among cognitively impaired participants, but estimates close to zero among cognitively unimpaired participants, while meta-regression showed a non-significant tendency towards larger effects in older samples. Collectively, these findings indicate that APOE-associated differences in BBB-linked fluid biomarkers may vary according to genotype, biomarker, and clinical context, while highlighting a potentially under-recognised association between APOE ε2 and barrier permeability (Bernocchi et al., 2026).

The most notable finding of this study is the increase of Q-Alb in APOE ε2 carriers. APOE ε2 is associated with reduced amyloid burden (Insel et al., 2021; Salvado et al., 2021), lower risk of late-onset AD (Belloy et al., 2023), and longevity (Shinohara et al., 2020).

However, emerging evidence indicates that ε2 is not uniformly protective. In particular, APOE ε2 has been associated with increased rates of Type 2 Cerebral Amyloid Angiopathy (CAA) (Love et al., 2014; Nelson et al., 2013; Yu et al., 2015), involving the deposition of amyloid peptides, including amyloid-β, in arterial vessels (Hu et al., 2025; Love et al., 2014). CAA is common in aging brains (Love et al., 2014), and is associated with an increased risk of intracerebral haemorrhage, particularly in ε2 carriers (Goldberg et al., 2020; Nicoll et al., 1997). CAA pathology reflects an imbalance between Aβ production and clearance, including via BBB-mediated clearance pathways (Hu et al., 2025). Indeed, CAA can both contribute to and worsen BBB dysfunction (del Valle et al., 2011; Magaki et al., 2018), and past research has demonstrated that CAA-affected vessels are more permeable to albumin (Wisniewski et al., 1997). Taken together, our findings of increased Q-Alb in ε2 carriers, and prior CAA research, implicate APOE ε2 in altered neurovascular and blood–CNS barrier function.

Bernocchi et al. demonstrated that this increased Q-Alb co-exists with favourable AD biomarker profiles, highlighting the need for further research into how APOE ε2-related barrier effects interact with its protective influence on AD risk (Bernocchi et al., 2026).

In contrast to APOE ε2, APOE ε4 carriage overall was not associated with significantly higher Q-Alb. There is substantial preclinical evidence demonstrating APOE ε4-mediated impairment of endothelial function, pericyte degeneration, neuroinflammation, and BBB disruption (Bell et al., 2012; Casey et al., 2015; Halliday et al., 2016; Jackson et al., 2022; Nishitsuji, K. et al., 2011; Yamazaki et al., 2020), as well as clinical research showing that the ε4 allele predisposes more severe Type 1 CAA, which involves capillary amyloid deposition with or without involvement of larger vessels (Love et al., 2014). Genotype-specific analyses provided some indication that ε4 effects may be obscured when ε4 carriers are considered as a single group. No effect on Q-Alb was observed in APOE ε3/ε4 carriers, the most common APOE ε4 genotype (Troutwine et al., 2022), and in mixed groups (which likely contained a substantial proportion of ε3/ε4 carriers given the population frequency of ε3). In comparison, medium, although non-significant and imprecise, elevations in Q-Alb were observed among ε4 homozygotes and ε2/ε4 carriers. This is consistent with the effects of the ε2 allele as discussed above, and with the established dose-dependent influence of APOE ε4 on dementia risk (Saddiki et al., 2020; Williams et al., 2026).

One possible explanation for the weak overall association is that Q-Alb may not capture the specific pattern of BBB dysfunction associated with APOE ε4. As an indicator of blood–CSF barrier function, Q-Alb provides an indirect measure of global blood–CNS barrier integrity and may be insensitive to subtle or regionally restricted BBB abnormalities (Hillmer et al., 2023). Neuroimaging studies have identified APOE ε4-associated BBB dysfunction within hippocampal and medial temporal regions (Montagne et al., 2020; Moon et al., 2021). Such focal alterations may have limited influence on Q-Alb, potentially contributing to the weak pooled association observed here.

The clinical composition of the Q-Alb literature may also have limited detection of genotype-specific effects. Most participants had established dementia, which is itself associated with elevated Q-Alb and BBB dysfunction (Llorens et al., 2015; Musaeus et al., 2020; Skillbäck et al., 2017; Wang et al., 2026; Wong et al., 2022). Consequently, the independent contribution of APOE ε4 may be difficult to distinguish in these samples. One study contributed a CJD subgroup (Karch et al., 2013), introducing additional clinical heterogeneity given its distinct pathophysiology (Noor et al., 2024; Watts et al., 2026).

However, Q-Alb did not differ significantly between CJD and other dementia groups in that study (Karch et al., 2013), the APOE ε4 effect was consistent with other diagnostic groups, and exclusion in sensitivity analysis did not materially alter the pooled estimates.

Consideration of diagnostic category in subgroup analysis did not provide evidence for a difference in APOE ε4 effects on Q-Alb across different dementia types (AD, non-AD dementia, and mixed samples). However, the limited representation of cognitively unimpaired participants prevented examination of whether APOE ε4–Q-Alb associations differ across the disease continuum.

By comparison, sPDGFRβ studies included participants ranging from cognitively unimpaired individuals to those with MCI or AD. APOE ε4 carriage showed a negligible association with sPDGFRβ overall, and examination of additional studies unable to be meta-analysed provided further support for the lack of association between APOE carrier status and sPDGFRβ. Effect estimates increased with mean participant age, although age was not a statistically significant moderator. Subgroup estimates also differed descriptively across diagnostic categories, with a small positive but non-significant effect among cognitively impaired participants and estimates close to zero among cognitively unimpaired participants. Narrative findings from Sagare et al (2026) provide convergent evidence for this pattern.

Studies examining sPDGFRβ alongside AD biomarkers suggest that the greater APOE ε4-associated pericyte injury and BBB dysfunction observed in cognitively impaired and AD populations may occur independently of established amyloid and tau pathology (Blumenfeld et al., 2024; Halliday et al., 2016; Montagne et al., 2020; Sagare et al., 2026).

### 4.1 Limitations and Future Directions

Several limitations of the available evidence should be acknowledged. The literature remains relatively small, particularly for sPDGFRβ and genotype-specific analyses, and many studies reported only aggregated APOE ε4 carrier status (Bowman et al., 2012; Edwards et al., 2025; Haghdel et al., 2024; Janelidze et al., 2017; Montagne et al., 2020; Motta et al., 2025; Preis, L. et al., 2024; Sagare et al., 2026; Tian et al., 2024), limiting assessment of genotype-specific effects. Several subgroup estimates were consequently imprecise, with wide confidence intervals indicating limited statistical power to detect genotype-specific associations. Considerable methodological and clinical heterogeneity was also present, including variation in age, sex, metabolic and vascular comorbidities, all of which may influence Q-Alb or sPDGFRβ (Brettschneider et al., 2005; Cicognola, Claudia et al., 2023; Janelidze et al., 2017; Parrado-Fernández et al., 2018; Preis, Lukas et al., 2024; Skillback et al., 2021; Tibbling et al., 1977). Finally, the predominantly cross-sectional evidence precludes determination of how APOE-associated changes in these biomarkers develop with ageing or disease progression.

Comparison of Q-Alb and sPDGFRβ findings was further limited by substantial differences in the populations contributing to each analysis. Q-Alb studies predominantly included participants with dementia, whereas sPDGFRβ studies largely comprised cognitively unimpaired participants. The limited sPDGFRβ literature also prevented genotype-specific meta-analyses, including examination of ε4 homozygotes. Finally, a key limitation of the sPDGFRβ meta-analysis was the inclusion of blood-based samples alongside CSF measures which, although less invasive, may be influenced by peripheral inflammation and vascular injury (Wang et al., 2026). Given potential biological differences between circulating and CSF sPDGFRβ, sensitivity analyses were conducted excluding one study with blood-based measurements and showed no material change to the pooled estimate.

Future research should prioritise large longitudinal cohorts with biomarker-defined disease classification, genotype-specific APOE reporting, and multimodal assessment of BBB and neurovascular integrity. In particular, measuring Q-Alb, sPDGFRβ, and other complementary BBB measures concurrently within the same participants would enable direct comparison of their relationships with APOE genotype across the disease continuum. Greater representation and consideration of APOE ε2 carriers is also warranted given the observed association with elevated Q-Alb and the possibility that ε2-associated alterations in barrier function arise through mechanisms distinct from the pericyte injury proposed for APOE ε4 (Blumenfeld et al., 2024). Such studies would help distinguish biomarker-specific effects from population differences and clarify the temporal and mechanistic relationships between APOE genotype and neurovascular dysfunction.

### 4.2 Conclusion

In conclusion, this meta-analysis found evidence that APOE ε2 carriage is associated with increased barrier permeability as measured by Q-Alb. In contrast, evidence for APOE ε4-related alterations in Q-Alb and sPDGFRβ was inconclusive: pooled estimates indicated negligible-to-moderate increases (consistent with greater BBB dysfunction) with exploratory analyses suggesting possible variation according to allele combination, diagnostic status, and age, although these estimates were imprecise and moderator effects were not statistically significant. These findings challenge the predominant focus on APOE ε4 in BBB research and suggest that APOE ε2 may also play an important, and potentially underrecognized, role in BBB disruption. More broadly, they indicate that APOE-related neurovascular effects are genotype-specific and warrant further investigation across the spectrum of cognitive ageing and dementia.

## Data Availability

All data produced in the present work are contained in the manuscript and supplementary files.

## Acknowledgements

The authors gratefully acknowledge the researchers who provided additional data and unpublished summary statistics in response to requests made during the conduct of this systematic review and meta-analysis.

## Declaration of generative AI and AI-assisted technologies in the manuscript preparation process

During the preparation of this work, the authors used ChatGPT to check the final manuscript for grammatical errors and inconsistencies. ChatGPT’s output consisted of a list of suggested revisions, which the authors reviewed and incorporated where appropriate. The authors take full responsibility for the content of the published article.

## Registration

DOI: 10.17605/OSF.IO/YX52P (OSF)

